# Healthcare workers benefit from second dose of COVID-19 mRNA vaccine: Effects of partial and full vaccination on sick leave duration and symptoms

**DOI:** 10.1101/2021.11.17.21266479

**Authors:** Earl Strum, Yolee Casagrande, Kim Newton, Jennifer B. Unger

## Abstract

**Importance:** In addition to morbidity and mortality of individuals, COVID-19 can affect staffing among organizations. It is important to determine whether vaccination can mitigate this burden. **Objective**: This study examined the association between COVID-19 vaccination status and time until return to work among 952 healthcare workers (HCW) who tested positive for COVID-19.

**Design:** Data were collected prospectively between December 2020 and July 2021. HCW who tested positive for COVID-19 completed an initial interview and were followed until they returned to work.

**Setting:** An academic campus in Southern California consisting of two large hospitals and multiple outpatient clinics and other facilities.

**Participants:** Clinical and nonclinical HCW who tested positive for COVID-19 during the study period (N=952, mean age=39.2 years, 69% female, 45% Hispanic, 14% white, 14% Asian/Pacific Islander, 5% African American, and 21% other race/ethnicity).

**Exposure:** COVID-19 vaccination status (unvaccinated, partially vaccinated, or fully vaccinated)

**Main Outcome Measures:** Days until return to work, presenting symptom

**Results:** Return-to-work time for fully vaccinated HCWs (mean=10.9 days) was significantly shorter than that of partially vaccinated HCWs (15.5 days), which in turn was significantly shorter than that of unvaccinated HCWs (18.0 days). Fully vaccinated HCWs also showed milder symptom profiles compared to partially vaccinated and unvaccinated HCWs.

**Conclusions and Relevance:** COVID-19 vaccination has the potential to prevent long absences from work and the adverse financial, staffing, and managerial consequences of these long absences.

**KEY POINTS:** *Question:* Do healthcare workers (HCW) who are vaccinated against COVID-19 return to work sooner and experience milder symptoms compared with unvaccinated HCW?

*Findings:* Among 952 healthcare workers who tested positive for COVID-19 between December 2020 and July 2021, return-to-work time for fully vaccinated HCWs (mean=10.9 days) was significantly shorter than that of partially vaccinated HCWs (15.5 days), which in turn was significantly shorter than that of unvaccinated HCWs (18.0 days). Fully vaccinated HCWs also showed milder symptom profiles compared to partially vaccinated and unvaccinated HCWs.

*Meaning:* COVID-19 vaccination has the potential to prevent long absences from work and the adverse financial, staffing, and managerial consequences of these long absences.

## INTRODUCTION

Research has documented that the Pfizer, Moderna, and Johnson & Johnson COVID-19 vaccines prevent hospitalization and mortality.^1^ However, few studies have examined the effects of COVID-19 vaccination on other nonclinical outcomes, including days missed from work. Although all HCWs who test positive for COVID-19 are advised to quarantine for 10 days, there is considerable variation in the number of days they actually spend recovering before returning to work.^2^ Focusing on return to work is important because long absences from work can cause adverse consequences for HCWs such as financial strain, unemployment, loss of healthcare, and mental health problems, as well as staffing and managerial challenges for organizations.

Several studies^3,4^ found that sick leave utilization among healthcare workers (HCW) declined significantly after vaccines became available. However, studies have not compared the sick leave utilization of vaccinated vs. unvaccinated employees. It is important to determine whether vaccinated HCW returned to work sooner quickly than unvaccinated or partially vaccinated workers during the same time in the history of the pandemic. This study compared the return-to-work time across three groups of HCWs who tested positive for COVID-19: those who were fully vaccinated with two Pfizer-BioNTech vaccine doses, those who were partially vaccinated with one dose, and those who were unvaccinated. We hypothesized that fully vaccinated HCWs would have shorter return-to-work times and milder symptoms than partially vaccinated HCWs, who in turn would have shorter return-to-work times and milder symptoms than unvaccinated HCWs.

## METHOD

The study was approved by the University of Southern California Institutional Review Board.

### Participants

The sample included clinical and nonclinical HCWs at an academic campus consisting of two large hospitals and multiple outpatient clinics and other facilities. Throughout the study period, December 17, 2020 to July 1, 2021, 4,686 HCWs were vaccinated with Pfizer-BioNTech vaccine. HCWs were tested for COVID-19 weekly or whenever they reported symptoms or exposures. HCWs who tested positive during the study period were eligible for inclusion in the study.

### Procedure

HCWs who tested positive for COVID-19 were contacted for an interview using a standardized questionnaire. Each COVID-19 positive HCW was followed longitudinally 3-4 times weekly by phone or text to assess symptoms until return-to-work criteria using medical center and CDC guidelines were met. The minimum quarantine period was 10 days with 24 hours of minimal to no symptoms prior to return-to-work clearance.

### Measures

The questionnaire included demographics, vaccination status, and the presenting symptom. Symptoms were grouped into categories: cough (cough, sputum), gastrointestinal (nausea, vomiting, diarrhea, anorexia, abdominal pain), shortness of breath, nasal (rhinorrhea, nasal congestion), fever/chills, throat (sore or itchy throat), loss of taste/smell, ocular symptoms (redness, burning, pain), chest pain, pain (headache/ myalgias), general (fatigue, weakness, dizziness).

*Return-to-work days* was the number of days between the HCW’s initial symptom onset or positive test date (whichever was earlier) and the first day back at work after recovering and/or quarantining. *Vaccination status* was defined as unvaccinated, partially vaccinated (> 3 days post first dose through day 13 post second dose) and fully vaccinated (> 13 days post dose 2). These data were obtained from the interviews and confirmed with medical records.

*Demographic covariates* included age, sex, and race/ethnicity.

### Data analysis

HCWs with return-to-work times > 100 days were excluded from the analytic sample because they represented special circumstances (e.g., employees who took extended leave because they were pregnant or caring for sick family members). Bivariable analyses (Analysis of Variance for continuous variables and chi-square for categorical variables) compared the three vaccination groups--unvaccinated, partially vaccinated, and fully vaccinated--on return-to-work days and demographics. Multiple regression analyses determined whether vaccination status was associated with return-to-work days, controlling for demographic variables. Chi-square analysis compared the three groups on COVID-19 symptoms experienced.

## RESULTS

### Descriptive statistics

A total of 1030 HCWs tested positive for COVID-19 during the study period and were therefore eligible for inclusion in the analysis; 78 were excluded because of extended leaves over 100 days, lack of follow-up data, or inconclusive vaccination status. This resulted in 952 COVID-19 positive HCWs: 811 unvaccinated, 116 partially vaccinated, and 25 vaccinated. Participants ranged in age from 19 to 71 years (mean=39.2, SD=11.1). The sample was 69% female, 45% Hispanic, 14% white, 14% Asian/Pacific Islander, 5% African American, and 21% other race/ethnicity. The mean return-to-work time was 18.49 days (SD=10.35).

### Univariate comparisons across the three vaccination groups

As shown in Table 1, the three groups did not differ significantly by age or sex, but fully vaccinated HCWs were less likely to be Hispanic (14% Hispanic) compared with partially vaccinated (44% Hispanic) and unvaccinated (46%) HCWs (p<.005).

**Table 1.** Univariate Comparison of HCWs who Tested Positive for COVID-19

|  | Fully Vaccinated | Partially Vaccinated | Unvaccinated | p-value |
| --- | --- | --- | --- | --- |
| N | 25 | 116 | 811 |  |
| Mean age (SD) | 43.2 (12.3) | 39.6 (11.4) | 39.0 (11.0) | .1665 |
| % Female | 76 | 68 | 68 | .6622 |
| % Hispanic | 14 | 44 | 46 | .0027 |
| % White | 24 | 16 | 13 | .2130 |
| % Asian/PI | 24 | 18 | 13 | .0745 |
| % Black | 47 | 4 | 6 | .7079 |
| Return-to-work days | 12.6 (2.9) | 16.5 (6.6) | 19.0 (10.8) | .0007 |

### Multivarible predictors of return-to-work days

Table 2 shows the results of the multiple regression analysis. After controlling for age, sex, and race/ethnicity, fully vaccinated HCWs (mean 10.9 days, β=-.111, p<.001) and partially vaccinated HCWs (mean=15.5 days, β=-.077, p<.05) had shorter return-to-work times, relative to unvaccinated HCWs (mean=18.0 days). Post hoc comparison of adjusted means revealed that fully vaccinated HCWs had shorter return-to-work times than partially vaccinated HCWs (t=2.01, p<.05). HCWs ages ≥40 had longer return-to-work times relative to younger HCWs. Sex and race/ethnicity were not significantly associated with return-to-work days.

**Table 2.**
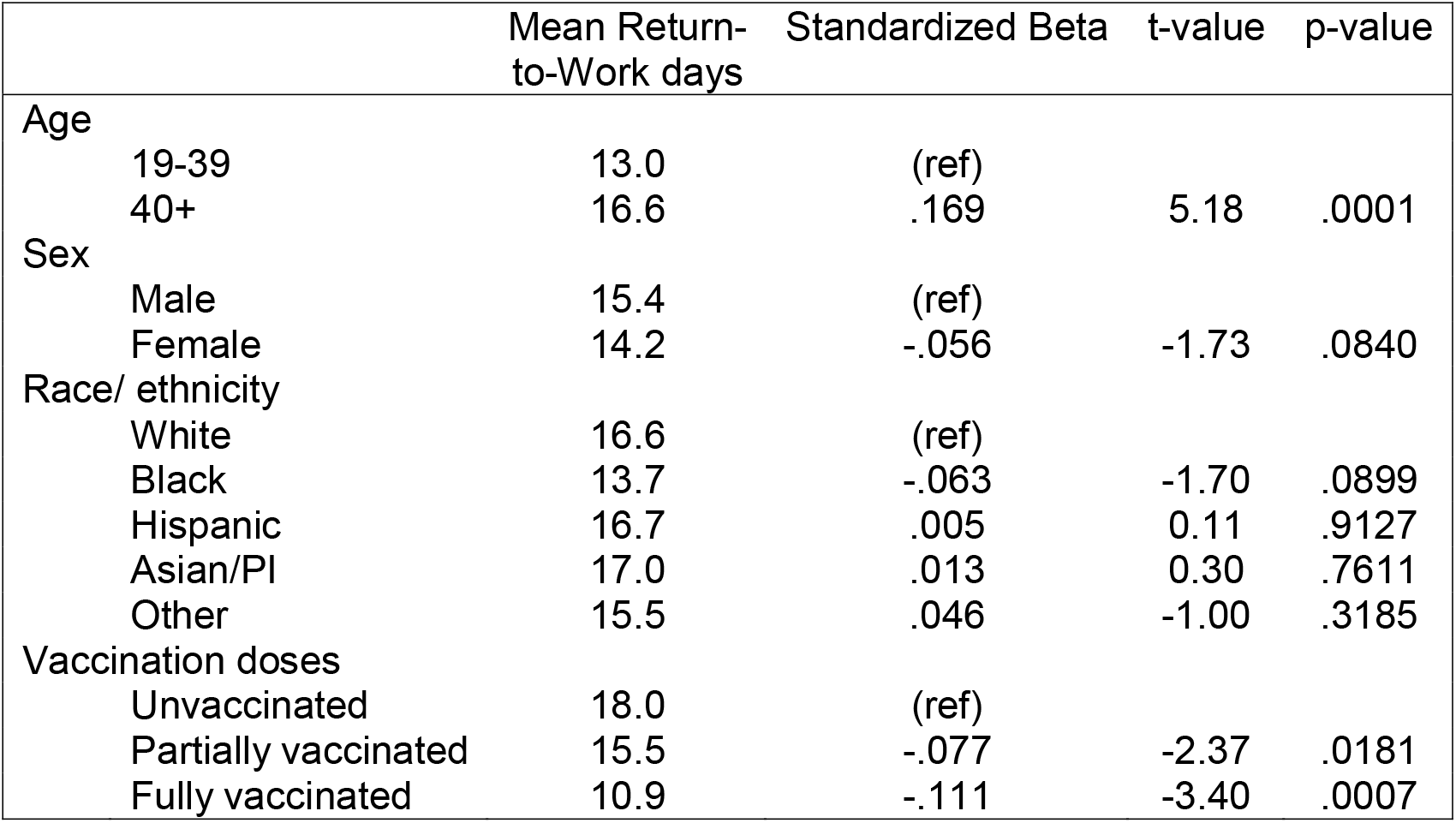
Variables Associated with Return-to-Work Days

|  | Mean Return-to-Work days | Standardized Beta | t-value | p-value |
| --- | --- | --- | --- | --- |
| Age |  |  |  |  |
| 19-39 | 13.0 | (ref) |  |  |
| 40+ | 16.6 | .169 | 5.18 | .0001 |
| Sex |  |  |  |  |
| Male | 15.4 | (ref) |  |  |
| Female | 14.2 | -.056 | -1.73 | .0840 |
| Race/ ethnicity |  |  |  |  |
| White | 16.6 | (ref) |  |  |
| Black | 13.7 | -.063 | -1.70 | .0899 |
| Hispanic | 16.7 | .005 | 0.11 | .9127 |
| Asian/PI | 17.0 | .013 | 0.30 | .7611 |
| Other | 15.5 | .046 | -1.00 | .3185 |
| Vaccination doses |  |  |  |  |
| Unvaccinated | 18.0 | (ref) |  |  |
| Partially vaccinated | 15.5 | -.077 | -2.37 | .0181 |
| Fully vaccinated | 10.9 | -.111 | -3.40 | .0007 |

### Presenting symptom

Figure 1 shows the presenting symptoms in each vaccination group. The most common presenting symptoms were asymptomatic (32%) and nasal (28%) among fully vaccinated HCWs; nasal (18%) and cough (16%) among partially vaccinated HCWs; and cough (15%) and throat (15%) among unvaccinated HCWs (chi-square=43.37, p=.0042).

**Figure 1.**
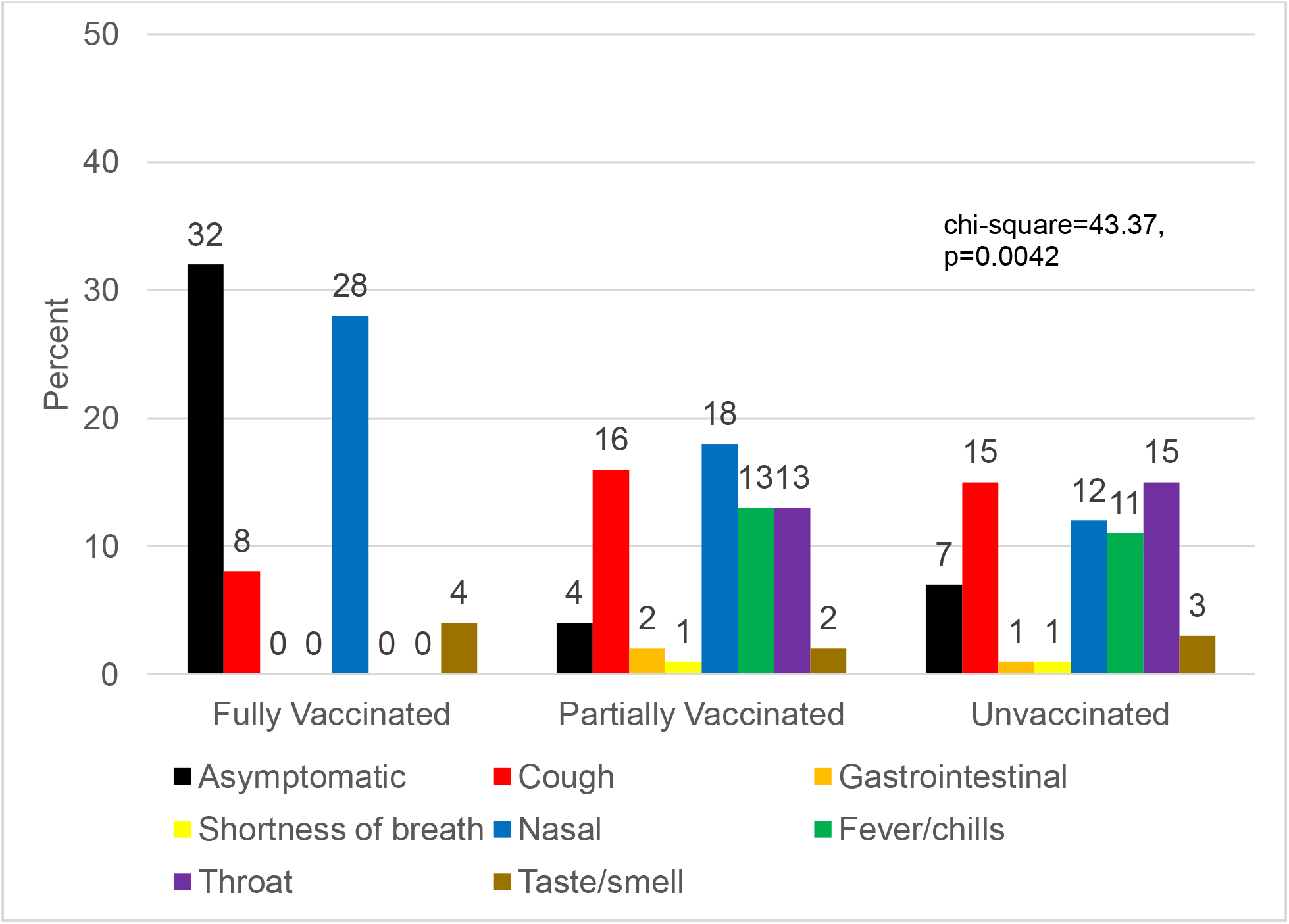
Presenting Symptom

## DISCUSSION

Fully vaccinated HCWs had fewer missed workdays and milder symptoms, compared to those who were incompletely vaccinated or unvaccinated. In addition to the health and financial benefits to the HCWs themselves, vaccination benefited the organization by returning HCWs to their jobs more quickly and reducing the need to hire temporary replacement staff who were likely less productive until they completed required training and learned institutional nuances. A reduced healthcare workforce can lead to increased costs based on overtime, premium shift differentials, closure of outpatient clinics and elective surgeries, and increased ambulance diversion resulting in reduced ability to accept critical admissions. Our findings demonstrate that COVID-19 vaccination benefits HCWs and healthcare organizations.

### Limitations

This study was conducted in early 2021, before the more virulent delta variant proliferated in the U.S.^5^ It is unclear whether the findings would be similar with the delta variant. Because of the increased risk of serious illness, hospitalization and transmission resulting from the Delta variant, had this study been conducted six months later, our results might have differed in the number of vaccine breakthroughs and possibly duration/severity of illness. It is also uncertain at this time whether waning immunity over time will increase return-to-work time among vaccinated HCWs. However we still believe that those who were fully vaccinated will still return to work more rapidly although the duration may be different and they will have a better overall outcome than those who are not vaccinated.

### Conclusion

These findings underscore the benefits of COVID-19 vaccination in reducing convalescent time and symptom severity, with fully vaccinated HCWs recovering more quickly and with a much greater likelihood of having no or milder symptoms. This has implications for public health and for a specific institution, both in terms of institutional costs and overall population health.

## Data Availability

All data produced in the present study are available upon reasonable request to the authors.

